# The COVID-19 Pandemic in Africa: Predictions using the SIR Model

**DOI:** 10.1101/2020.06.01.20118893

**Authors:** Musalula Sinkala, Panji Nkhoma, Mildred Zulu, Doris Kafita, Rabecca Tembo, Victor Daka

**Affiliations:** University of Zambia, School of Health Sciences, Department of Biomedical Sciences, P.O. Box 50110, Nationalist Road, Lusaka, Zambia; University of Zambia, School of Medicine, Department of Pathology and Microbiology, P.O. Box 50110, Nationalist Road, Lusaka, Zambia; Copperbelt University, School of Medicine, Ndola, Zambia

**Keywords:** COVID-19, COVID-19 in Africa, SIR model, Disease prediction, COVID-19 modelling

## Abstract

Since the earliest reports of the Coronavirus disease - 2019 (COVID-19) in Wuhan, China in December 2019, the disease has rapidly spread worldwide, attaining pandemic levels in early March 2020. However, the spread of COVID-19 has differed in the African setting compared to countries on other continents. To predict the spread of COVID-19 in Africa and within each country on the continent, we applied a Susceptible-Infectious-Recovered mathematical model. Here, our results show that, overall, Africa is currently (July 24, 2020) at the peak of the COVID-19 pandemic, after which we predict the number of cases would begin to fall in August 2020. Furthermore, we predict that the ending phase of the pandemic would be in Late-November 2020 and that decreasing cases of COVID-19 infections would be detected until around August 2021 and September 2021. Our results also reveal that of the 51 countries with reported COVID-19 cases, only eight, including Algeria, Morocco and Zambia, are likely to report higher monthly COVID-19 cases in the coming months of 2020 than those reported in the previous months. Overall, at the end of this pandemic, we predict that approximately 2,201,849 (about 1,451,567 future cases) individuals in Africa would have been infected with the COVID-19 virus. Here, our predictions are data-driven and based on the previously observed trends in the spread of the COVID-19 pandemic. Shifts in the population dynamics and/or changes in the infectiousness of the COVID-19 virus may require new forecasts of the disease spread.

## Introduction

The novel Coronavirus disease-2019 (COVID-19), which was first reported in China in December 2019, has quickly spread to become a global pandemic [1–3]. As of 24^th^ July 2020, over 15.8 million people have tested positive for COVID-19 [4]. So far, global infections of the virus have been unevenly distributed across continents and countries; Europe, North America and South America are among the most impacted. Despite more than 750,282 cases reported in Africa, the spread of COVID-19 has been surprisingly slow, and the disease has exhibited lower-case fatality rates in comparison to other continents [5]. It was expected that the continent, with fragile health systems, barriers to testing, and potentially vulnerable populations, would report high numbers of cases and deaths. Additionally, familiarity with infectious disease outbreaks and diseases leading to an educated immune system has been postulated as a possible reason for these observations [5,6].

Recently, mathematical models have been applied to investigate the spread of the COVID-19 pandemic in various countries, among others, China [7], Italy [8], and England [9]. The predictions gleaned from these models have offered a platform for decision making aimed at controlling and/or mitigating the spread of COVID-19 pandemic and the optimisation of lockdowns and treatment efforts [9–11]. However, most of the recent COVID-19 modelling has been in high-income countries, and very few efforts have been made to model the spread of COVID-19 in many African countries.

Mathematical models that have been employed to predict the spread of the pandemic include logistic models [12,13] and Susceptible-Infected-Recovered (SIR) models [8,14]. In modelling using the SIR approach, we assume that the population is a compartment of interacting individuals in which the disease spread from the infected to the susceptible, and the infected either recover and build an immunity toward the infectious agent or succumb to the infection [15,16]. Here, we use the SIR model to predict the spread of the COVID-19 positive cases on the African continent as a whole and independently for each of the 45 different countries on the continent. Overall, we provide valuable intuition regarding the expected trajectory of the disease in Africa.

## Results

### COVID-19 in Africa

We obtained a dataset of the global COVID-19 cases from covid.ourworldindata.org. We found that out, as of July 24, 2020, 45 out of 56 African counties have reported the number of COVID-19 positive cases.

We extracted information on COVID-19 cases in African countries to show that South Africa has reported the highest number of cases (408,052), followed by Egypt (90,413) and Nigeria (38,948; Figure 1).

**Figure 1:**
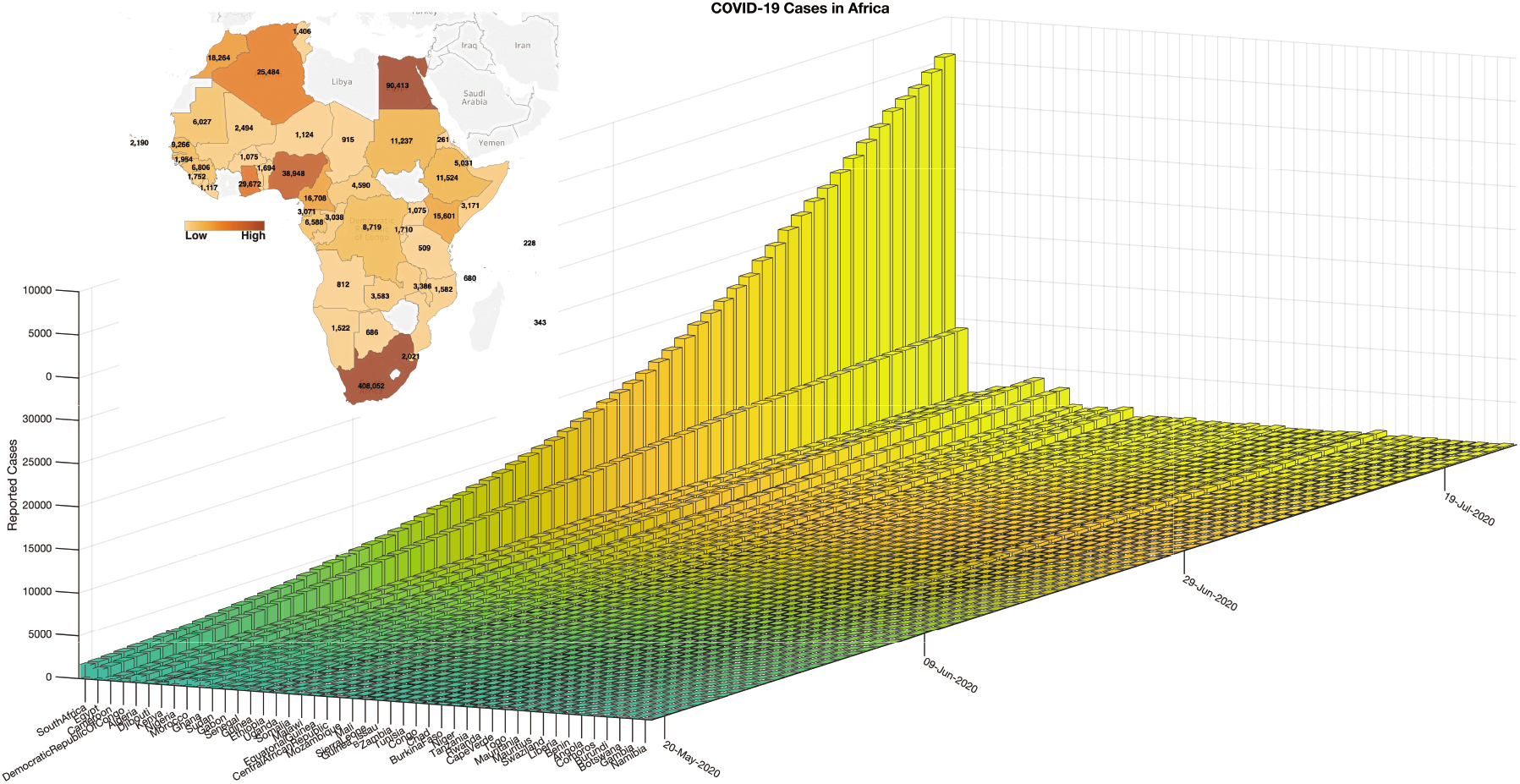
Status of the COVID-19 pandemic in Africa as of 28th May 2020

### Prediction of COVID-19 cases in Africa

We obtained the daily COVID-19 cases in Africa starting March 1, 2020, by aggregating all the reported cases per day for each country in Africa. Then, we utilised the susceptible-infected-removed (SIR) epidemic model to estimate the trajectory of the pandemic in Africa (see Methods Sections). Here, our model has accurately predicted the cases of COVID-19 in Africa with statistical significance (R^2^ = 0.999, p-value = 1.3 × 10^−253^; Figure 2). Furthermore, our results showed that, in Africa, the COVID-19 pandemic would be at the turning point on August 8 2020, get to the steady growth phase around October 2 2020, and the ending phase will begin around November 24 2020. Overall, based on the current data and the trajectory of COVID-19 positive cases, we predict that COVID-19 will infect another 1,451,567 individuals to bring the total number of COVID-19 positive cases to about 2,201,849. Also, we predict that the pandemic would have ended on the dates between August 2021 and September 2021 (for more details, see Table 1).

**Table 1:** Prediction of COVID-19 spread in Africa by SIR model

| Parameter | Statistic |
| --- | --- |
| Estimated duration (days) |  |
| Turning day | 160 |
| Acceleration phase | 51 days |
| Deceleration phase | 55 days |
| Total growth phase duration | 106 days |
| Total epidemic duration | 577 days |
| Estimated datums |  |
| Outbreak | 01-Mar-2020 |
| Start of acceleration | 17-Jun-2020 |
| Turning point | 08-Aug-2020 |
| Start of steady growth | 02-Oct-2020 |
| Start of ending phase | 24-Nov-2020 |
| End of the epidemic (5 cases) | 14-Aug-2021 |
| End of the epidemic (1 case) | 29-Sep-2021 |
| Statistics (total cases) |  |
| F-statistics vs zero model | 178,073 |
| p-value | $1.31 \times 10^{-253}$ |

**Figure 2:**
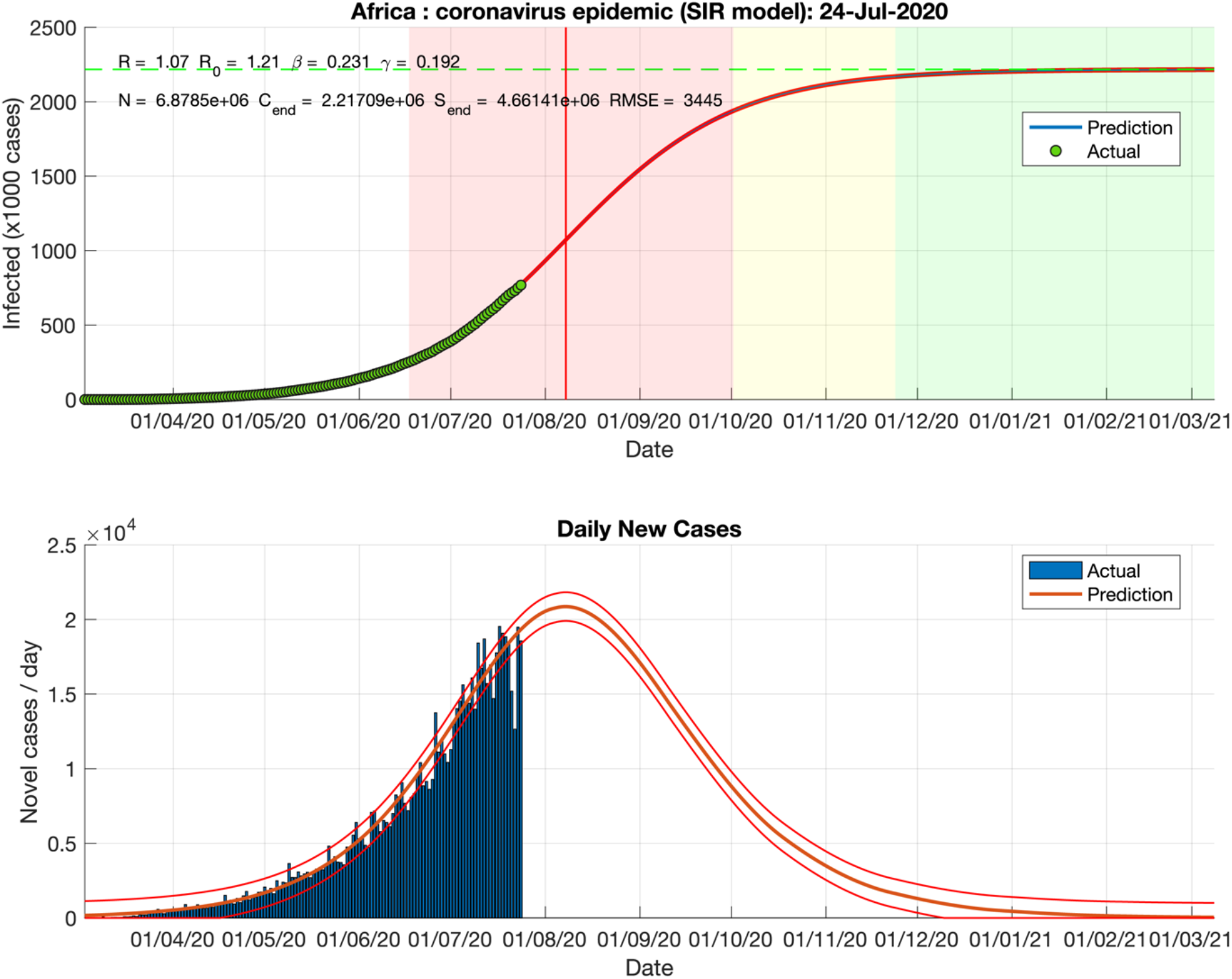
Predicted and actual cases of COVID-19 based on the SIR model as of 28 May 2020.

### Prediction of COVID-19 cases for each African country

Next, we used the SIR model to predict the spread of COVID-19, the expected total number infected, and the duration of an epidemic in each African Country. Here, our results showed that, for most countries in Africa, we would expect to see fewer monthly COVID-19 cases from September 2020 than those reported in the previous months (Figure 3a; see Supplementary File 1 for all predictions, also see the individual plots for each country provided in Supplementary File 2). However, based on the current data and spread of the disease, we predicted that the cases of COVID-19 would likely peak in July 2020 for Nigeria (Figure 3b), and in August 2020 for South Africa (Figure 3c), and among other countries. Furthermore, we predict the number of COVID-19 cases in Morocco, Kenya and among other countries to peak in August.

**Figure 3:**
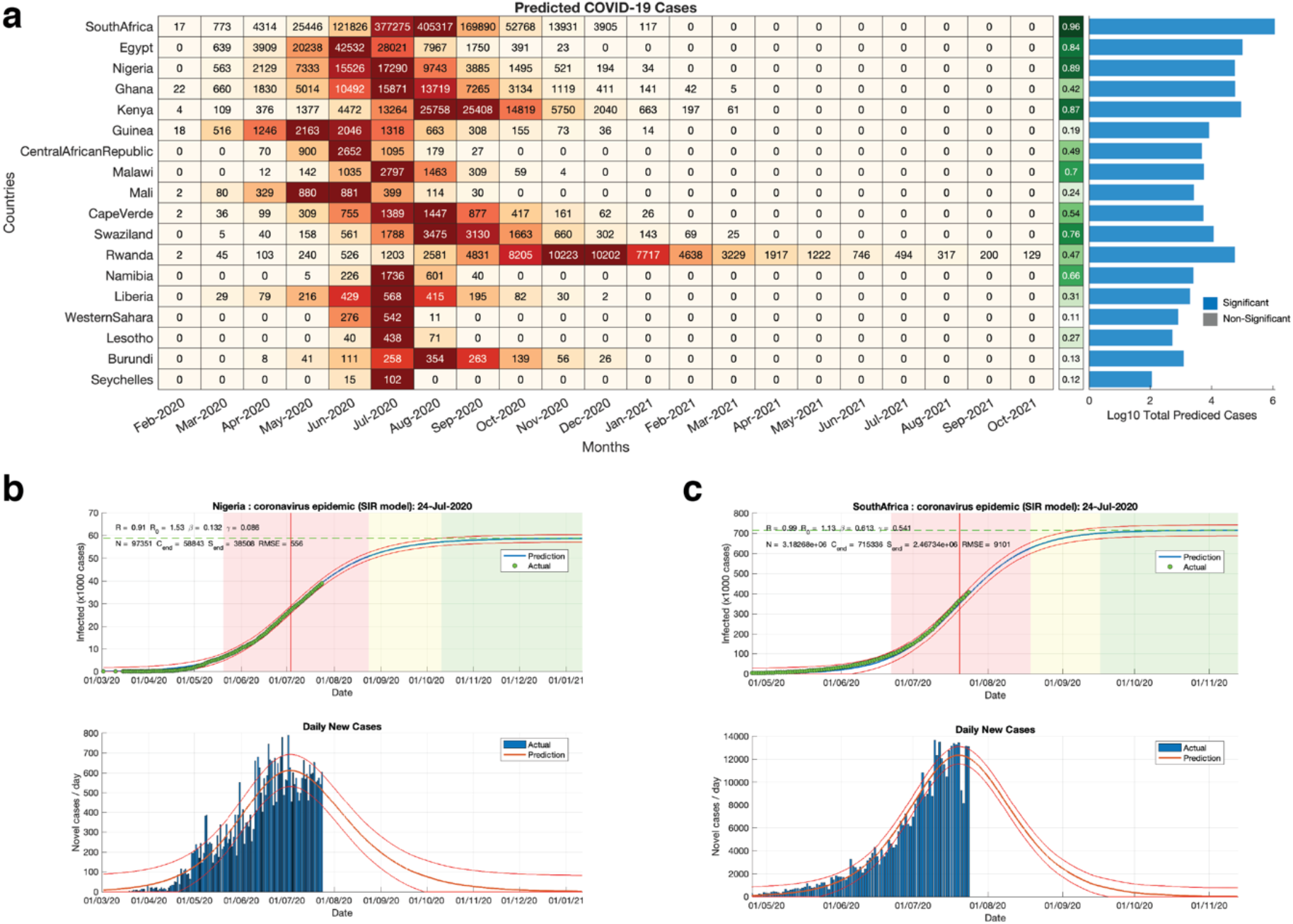
**(a)** Left; heatmap showing the predicted COVID-19 cases per month based on the SIR model for each country in Africa. Middle; Pearson’s linear correlation (R-square) between the predicted and actual daily COVID-19 across each country. The bar graphs represent the based ten logarithms transformed total numbers of predicted COVID-19 cases from the start to the end of the COVID-19 pandemic. Note: the strength of the predictions made for each country should be evaluated based on the given R-square values. Other African countries are not shown in the heatmap because our SIR-model had failed to make statistically significant predictions. The bars are coloured based on the false discovery rate p-values of the SIR model of each country; blue denotes q-values < 0.05 and grey denotes q-values > 0.05. Predicted and actual cases of COVID-19 based on the SIR model in **(b)** Egypt and **(c)** South Africa as of 24 July May 2020.

### COVID-19 in most affected African countries versus the rest of the World

We compared the reported COVID-19 positive cases in the most impacted countries in Africa (South Africa and Egypt) to those in other countries that have been affected across the globe. Here, we showed that the scale of the COVID-19 cases has steadily increased, with the scale of the pandemic in South Africa supposing that in Japan, China, Italy, and United Kingdom (Figure 4a). This assertion remains valid even when we consider the number of reported COVID-19 cases as a per cent for each of the country’s population size (Figure 4b).

**Figure 4:**
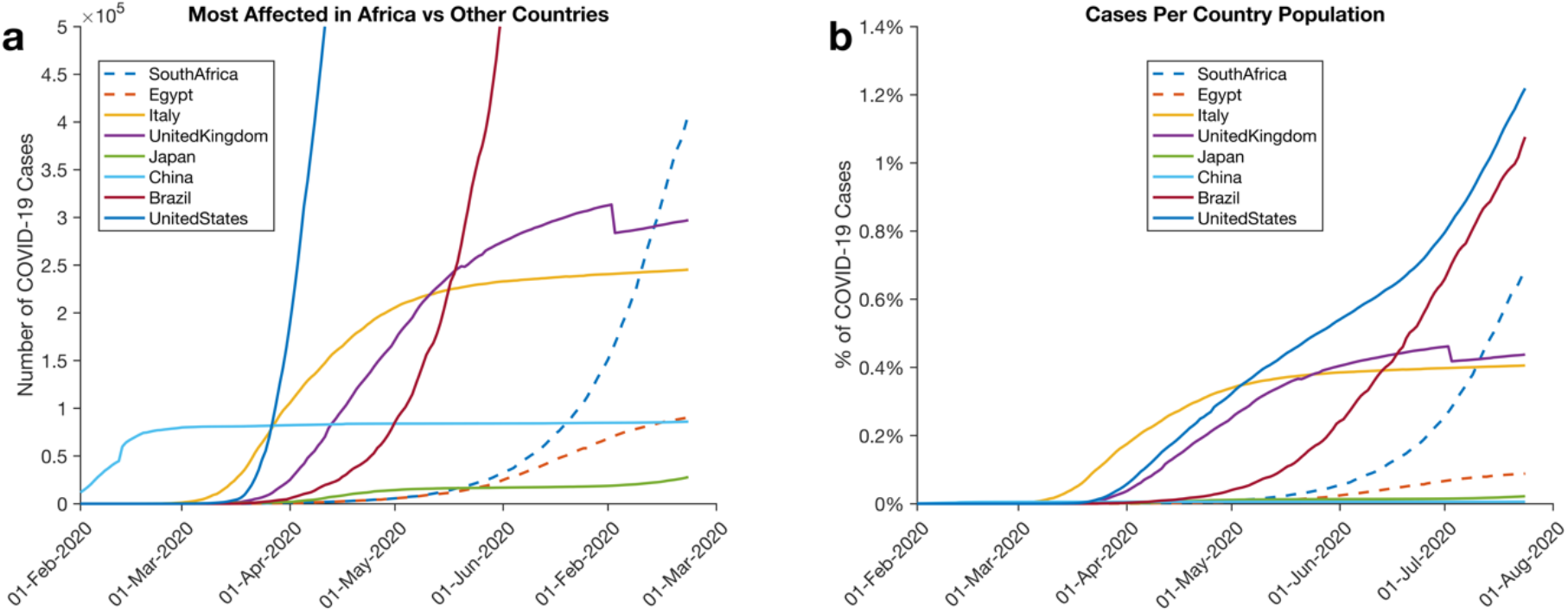
Comparison of COVID-19 Cases reported in some of the most affected countries in Africa (South Africa and Egypt) and countries on other continents. **(a)** Lines show the total number of reported COVID-19 cases. **(b)** Lines show the percentage of COVID-19 infected individuals for each country.

## Discussion

We conducted a predictive analysis of the COVID-19 pandemic in Africa. Our results show that the scale of the pandemic is low across many countries in Africa. Many experts have debated the reason why fewer cases of COVID-19 are being reported in Africa. Some have pointed toward the lack of widespread testing of COVID-19 [17–20], whereas others point towards the African climate [21,22].

We showed that, on average, most counties in Africa, including Zimbabwe, Malawi, and Benin, have reported fewer than 10,000 COVID-19 cases. Conversely, most European countries have reported, on average more than 30,000 COVID-19 cases. Importantly, in most of the counties on the continent, our prediction shows that the number of COVID-19 cases reported in July 2020 are unlikely to be surpassed by those in the next few months. Furthermore, we predicted that the COVID-19 pandemic would disproportionately affect different countries in Africa. We expect that even the worst affected among these would report COVID-19 positive cases that are comparable to those other regions of the World.

Interestingly, our model predictions were statistically significantly valid for only 18 of the 51 individual African countries for which we predicted the spread of the COVID-19 pandemics. Here, our model provided wrong predictions of the COVID-19 pandemic in 33 different African countries. We noted that in all these cases, our model failed because of the inconsistent reporting of the COVID-19 cases. One such case among the 33 counties is Cameroon: which on the 6^th^ of June 2020 had reported 810 new COVID-19 infections and no new COVID-19 infections on 7^th^ June 2020. Furthermore, Cameroon reported no new COVID-19 cases between the 27^th^ of June to the 5^th^ of July but reported more than 1,100 new COVID-19 cases on the 6^th^ of July 2020. Therefore, our results imply that efforts to model the spread of COVID-19 in many African countries are significantly hampered by the dependability of the currently available COVID-19 cases data.

Altogether, based on the current COVID-19 pandemic data and the spread of disease, our predictions show that the peak of the pandemic is likely to be in August 2020 for most African countries. Here, we caution that our results are only as good as the data and the previous trend observed in the spread of the COVID-19 pandemic. Hence, these predictions may not be indicative of future trends where certain parameters, such as the population dynamics and COVID-19 virus infectiousness, are shifted from those currently prevailing. Therefore, we encourage everyone (including individuals in African countries) to adhere to the guidelines that are aimed at reducing the spread of the COVID-19 virus as provided by the WHO and other relevant organisations.

## Methods

We analysed a COVID-19 dataset representing the reported virus-positive cases in the World as of July 24, 2020, obtained from https://covid.ourworldindata.org/data. We extracted the COVID-19 cases reported in 51 African countries and plotted these individually to show the trajectory of the COVID-19 pandemic for each country.

Next, we aggregated all the reported COVID-19 cases in Africa since March 1, 2020. Then we used the Susceptible-Infected-Removed (SIR) mathematical model [15,16] to predict the temporal dynamics of the COVID-19 spread, the expected number of infection cases, and the duration of the pandemic in Africa. Briefly, the principal assumption of the SIR model is that the population, in which the viruses (or other pathogens) spreads is comprised of three subgroups of individuals. Those uninfected and susceptible (S) to infection, 2) those infected (I) and can transmit the infection to the uninfected, 3) and those individuals removed (R) from the infection cycle, either because they recovered from the disease and are immune or succumbed to the disease [23,24] [10]. Here, we applied the SIR model implemented in MATLAB [14,25] that we have deposited here: https://www.mathworks.com/matlabcentral/fileexchange/76435-covid-19-predictions-in-africa-using-the-sir-model.

Furthermore, we used the SIR model to predict the spread of COVID-19, the expected number of infection cases, and the duration of the pandemic for each country in Africa. Here, we modified the code also to predict the number of COVID-19 infected cases of each country per day (see the Code Availability sections).

Briefly, we obtained the predicted number of COVID-19 using the SIR model. These predictions include multiple data points per day. Therefore, we aggregated the predictions made at irregular time intervals per day to yield a single prediction for each country per day by interpolation using the Cubic spline algorithm [26,27]. Also, to predict the number of cases of COVID-19 infection of each country, we aggregated the predictions for each month (see Figure 3a).

## Data Availability

The data that support the findings of this study are available at covid.ourworldindata.org.

## Data Availability

The data that support the findings of this study are available at covid.ourworldindata.org.

## Code Availability

We have deposited all the MATLAB source code used to process, analyse, and reproduce the major finding of this report at the following websites: https://www.mathworks.com/matlabcentral/fileexchange/76435-covid-19-predictions-in-africa-using-the-sir-model

## Supplemental Information

**Supplementary File 1:** Predict cases of COVID-19 using the SIR model for each day across each country in Africa.

**Supplementary File 2:** Plots of the predicted trajectory of the COVID-19 cases of each country in Africa.

## Author Contributions

SM, PN, and VD conceptualised the study; The formal methodology was devised by VD, DK, PN, MZ, RT, and MS; VD, DK, PN, RT, and MS performed the formal analysis of the datasets; VD, DK, MZ, RT, and PN wrote the draft manuscript; the manuscript was revised by MS, DK, MZ, RT, VD. MS created visualisations.

